# Implementing an EHR-Integrated Quality and Safety Dashboard to Improve Inpatient Safety: Clinical Usage and Preliminary Outcomes

**DOI:** 10.64898/2026.10.26.26366493

**Authors:** Marc Philip Pimentel, Savanna Plombon, Nicholas Piniella, Alison Garber, Maria A Malik, Pamela Garabedian, Erin O’Fallon, Catherine Yoon, Julie Fiskio, Stuart Lipsitz, Jeffrey L Schnipper, Anuj K Dalal

## Abstract

**Background:** Data driven tools are increasingly used to support patient safety and quality improvement, yet evidence on the impact of integrated quality and safety dashboards remains limited.

**Methods:** We conducted a pre–post implementation study using interrupted time series analysis to evaluate the impact of an EHR-integrated quality and safety dashboard on clinical processes and patient outcomes, among three general medicine units at a 171-bed community teaching hospital. All adult patients admitted during seven-month pre- and post-implementation periods were included. The primary outcome was the proportion of hospitalizations with at least one adverse event. Secondary and exploratory outcomes included hospital-acquired conditions, adherence to safety-related process measures, and dashboard utilization.

**Results:** A total of 9,023 hospitalizations were included (4,640 pre-implementation; 4,383 post**-**implementation). The proportion of hospitalizations with at least one adverse event was similar pre-implementation (23.15%; 95% CI, 20.96–25.29) and post-implementation (22.97%; 95% CI, 20.86–25.23). CLABSI per 100 patient-days was reduced (aOR 0.47, 95% CI [0.37-0.60]); and exploratory analyses demonstrated decreases in slope for post-discharge venous thromboembolism (aOR, 0.66; 95% CI, 0.48–0.91; *p*=0.01) and hospital-acquired pressure injury (aOR, 0.69; 95% CI, 0.48–1.00; *p*=0.05). Several process measures improved, including code status confirmation, venous thromboembolism prophylaxis, delirium screening, chlorhexidine bathing, and fall precautions.

**Conclusions:** Implementation of an EHR-integrated quality and safety dashboard was associated with meaningful improvements in multiple care processes and CLABSI but not overall adverse events. These findings highlight the potential of workflow-integrated dashboards to support frontline quality improvement, while underscoring the need for additional measures to improve clinical outcomes.

## INTRODUCTION

Despite widespread adoption of electronic health records (EHRs), hospitals continue to struggle with implementing best practices to prevent hospital-acquired conditions (HACs) and other sources of patient harm.^1–5^ As hospitals move towards implementing High Reliability Organization (HRO) models for quality improvement, the concepts of engaging staff with key quality measures, while providing robust support for information technology solutions, become critical for successful implementation.^6^ While EHRs enable near-real time access to granular clinical data (e.g., vital signs, medication administration, fluid balance) that are essential for frontline clinicians to adhere to best practices, long-standing patient safety risks have persisted– and in some cases worsened–since widespread EHR adoption.^7, 8^ Adverse events (AEs) among hospitalized patients remain common; a recent study by Bates et al. found that nearly 1 in 4 inpatients experience an AE, with 23% deemed to be preventable^9.^

One contributing factor is the fragmented design of most EHR interfaces, which increases cognitive burden and impairs clinicians’ ability to synthesize relevant data and provide guideline-concordant care.^10–15^ For instance, the data needed to assess risk for HACs such as catheter-associated urinary tract infections (CAUTI) are often scattered across different EHR tabs and flowsheets–catheter-days in flowsheet, indications in orders, urine culture results in labs–hindering frontline clinicians from synthesizing information into knowledge to drive clinically relevant actions, such as early removal of indwelling urinary catheters.

Beyond these user design barriers, quality and safety leaders face organizational challenges in mobilizing frontline clinicians around healthcare improvement efforts. Physicians, advanced practice providers, and nurses often report to different departments and service lines, thereby making communication of institutional priorities challenging.^16^ Physicians are frequently interested in quality improvement work, but many lack the time to regularly engage and execute on the work. Ideally, inter-professional care teams using HRO models would have regular interactions with quality measures pertinent to their unit, but on a day-to-day basis frontline staff may lack access to, or struggle to interpret, abstract quality improvement measures (e.g., device-utilization ratios or hospital-acquired infection surveillance measures), which are typically designed for administrators and analytics teams.^17–18^

To address these gaps, we designed and developed a customized, EHR-integrated dashboard to support real-time identification and mitigation of hospital safety risks at the patient, unit, and service level during our institution’s transition to a new commercial EHR system.^19–20 6^ Due to governance constraints and technical limitations during the EHR roll-out, we optimized this dashboard within the context of an ongoing patient safety research program.^17^ The dashboard was tailored to align with institutional quality priorities and frontline clinician workflows, allowing users to quickly identify patients not meeting key safety process measures and take immediate, guideline-aligned actions, such as placing orders or discontinuing devices.^21–25^ In this quality improvement study, we evaluated the effect of implementing this custom-developed EHR-integrated dashboard on process and outcome measures related to HACs at an academically affiliated community hospital. We also assessed adoption and usage during the implementation period to better understand real-world integration into clinical workflows.

## METHODS

### Overview of Research Questions and Methods by Implementation Domain

We used the Consolidated Framework for Implementation Research (CFIR) to guide the design, deployment, and evaluation of our custom-developed dashboard.^30^ As part of prior efforts,^22–24 26–28^ we identified relevant research questions and methods (**Table 1**) across five CFIR domains: innovation, outer setting, inner setting, individual roles and characteristics, and implementation process.^29–30^ We refined the initial user requirements for the innovation that were identified through focus groups and interviews and incorporated into the initial version of the dashboard, which we had implemented at a tertiary academic medical center.^21–25, 27–28^ For the current study, we iteratively refined these requirements based on shifts in the external policy environment (e.g., Centers for Medicare and Medicaid [CMS] Hospital-Acquired Condition Reduction Program), evolving institutional quality and safety priorities aligned with these external mandates, and workflow constraints and usability feedback from frontline clinicians working in a community hospital setting.^26^ This iterative, context-sensitive development process was essential to ensuring alignment between the dashboard’s functionality and real-world clinical needs. By tailoring the innovation to both organizational goals and frontline realities, we aimed to support adoption, integration into existing workflows, and sustained use across clinical teams at the community hospital. The Mass General Brigham (MGB) Human Research Committee approved this study.

**Table 1.** Application of CFIR* Domains to Research Questions and Corresponding Methods.

| CFIR Domain and Construct | Research Question | Methods* |
| --- | --- | --- |
| Intervention<br>Characteristics:<br>Intervention Source: | Which EHR data correspond to institutionally defined quality measures and are routinely updated during clinical care? | Conventional content analysis of feedback from interviews and focus groups with stakeholders to refine requirements |
| Outer Setting: External Policies and Incentives | What outcome measures have mandatory public reporting and pay-for-performance requirements that clinicians should know about? | Meetings with quality and safety leadership and clinical service leaders to align key outcomes |
| Inner Setting:<br>Implementation Climate:<br>Relative Priority,<br>Compatibility | What local quality improvement initiatives related to publicly reported outcome measures should clinicians know about? | Review of hospital quality improvement initiatives and metrics |
|  | Which process and outcome measures are important to the institution and meaningful to clinicians? | Selection of relevant process and outcome measures |
|  | Which workflows are most acceptable for clinicians to access on a dashboard? | Focus groups with nurse managers, unit leadership, frontline clinicians |
| Individuals – Roles and Characteristics: Individual Stage of Change | Which frontline clinicians will use the dashboard? | Review of dashboard usage reports and metrics |
| Implementation Process Domain: Planning | What are the barriers to incorporating a dashboard into clinical workflows? | Focus groups with frontline clinicians and clinical leaders. |
| *Consolidated Framework for Implementation Research (Original Constructs). Some of these methods / studies were conducted as part of prior or concurrent studies |  |  |

### Study Design, Setting, Participants

This pre-post implementation study was conducted on the general medicine units at Brigham and Women’s Faulkner Hospital (BWFH), a 171-bed community teaching hospital affiliated with MGB in Boston, MA. The three general medicine units were staffed by attending hospitalists, advanced practice providers (APPs) and internal medicine residents. All units conducted weekday interdisciplinary rounds (IDRs), which included physicians, APPs, nurses, care coordinators, social workers, physical therapists, and nutritionists. During IDRs, teams routinely reviewed quality improvement data relevant to the unit–such as code status documentation, central line care, and urinary catheter use. All clinicians used a shared commercial EHR (Epic Systems, Inc., version 2018), standardized across MGB hospitals.

All adult patients (aged ≥18 years) admitted to the three general medicine units during the seven-month pre-implementation (9/2018 to 3/2019) and post-implementation (9/2019 to 3/2020) periods were included. A six-month wash-in period (3/2019 to 8/2019) was used to formally introduce the dashboard, support integration into clinical workflows, and account for seasonal changes in patient populations and trainee experience. There were no exclusion criteria. The exposures associated with pre-implementation (usual care) and post-implementation (integration of dashboard into clinical workflows) are illustrated in **Figure 1**.

**Figure 1.**
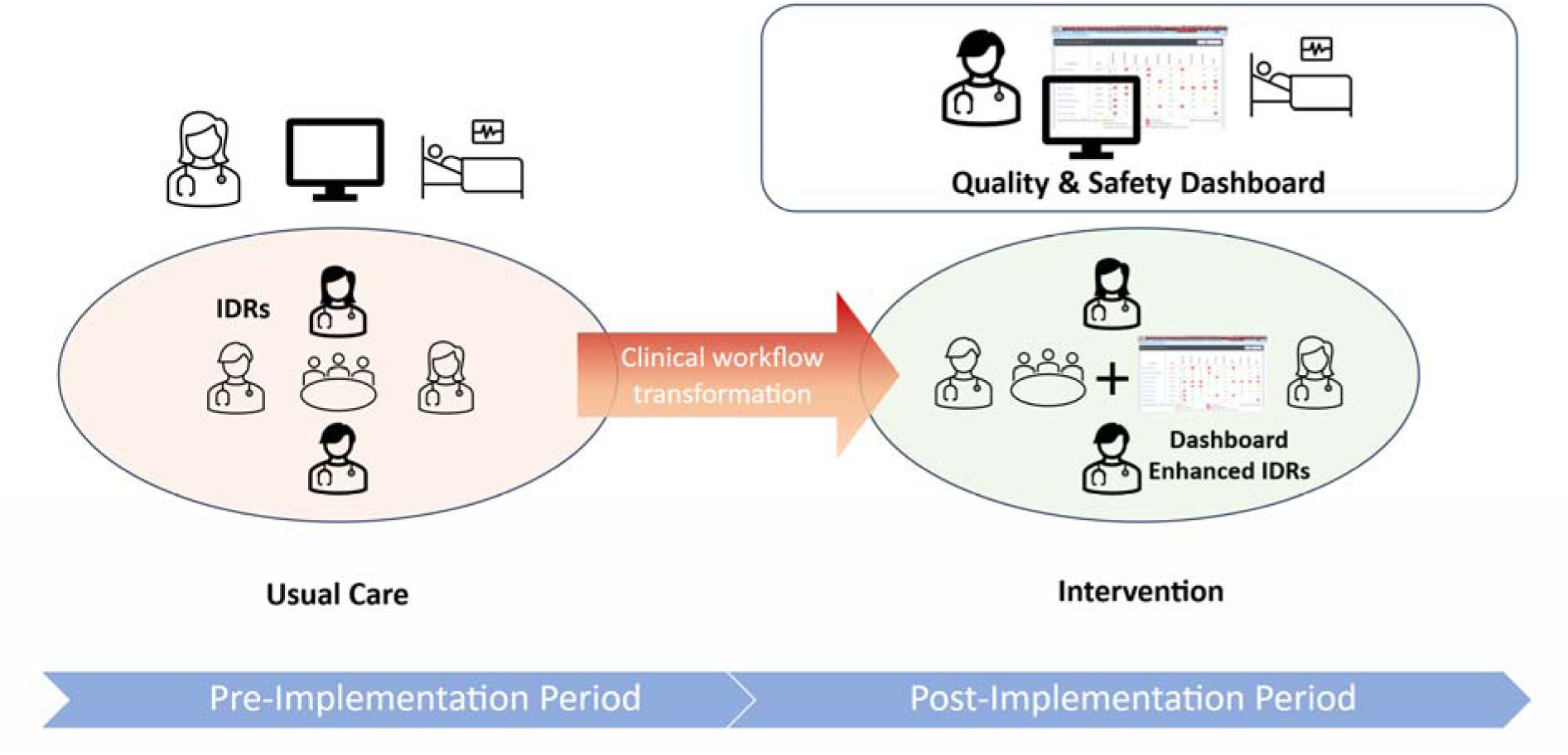
Incorporating a Quality and Safety Dashboard into Clinical Workflows

### Overview of Quality & Safety Dashboard

The clinician-facing dashboard (**Figure 2**) was developed, refined, and integrated into the EHR through an iterative user-centered design process previously described. ^22–24 26–28^ Its primary aim was to support frontline clinicians in identifying patients with common hospital-related safety risks and to promote adherence to institutional current and emerging quality and safety guidelines (**Table 2**). Hypoglycemia was tracked as a control measure because it was not in the dashboard during the study period.

**Figure 2.**
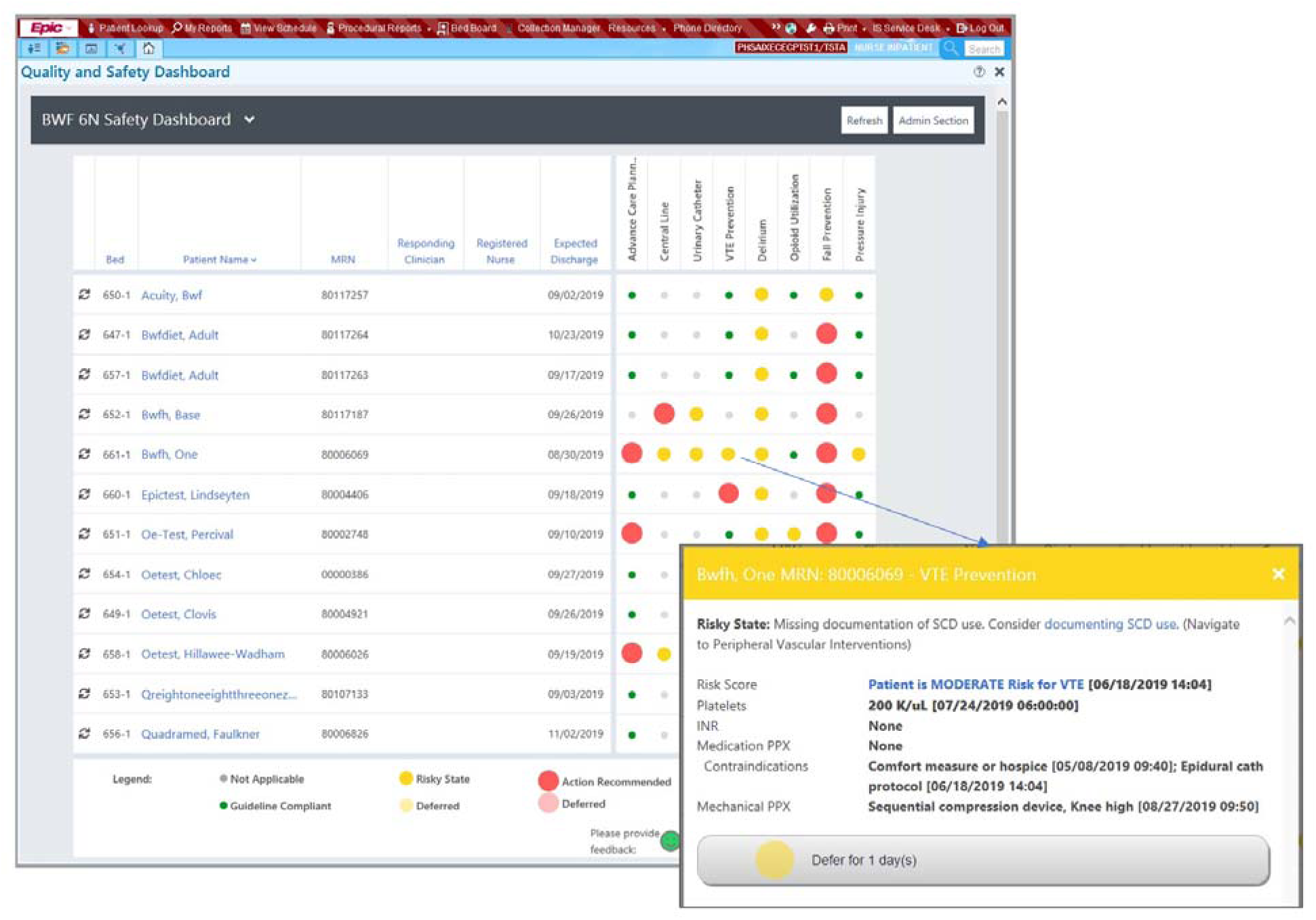
Screen shots of Quality and Safety Dashboard. The interactive graphical user interface supports frontline clinician workflow through unit and team-based patient views and domain-specific patient risk flags. Key usability enhancements included customizable patient lists, embedded patient chart views displaying risk flags within the EHR, actionable hyperlinks enabling real-time clinical actions, flexible sorting options, configurable notifications for newly triggered flags, transparent logic definitions explaining risk indicators, and ‘defer’ functionality to reduce alert fatigue.

**Table 2.**
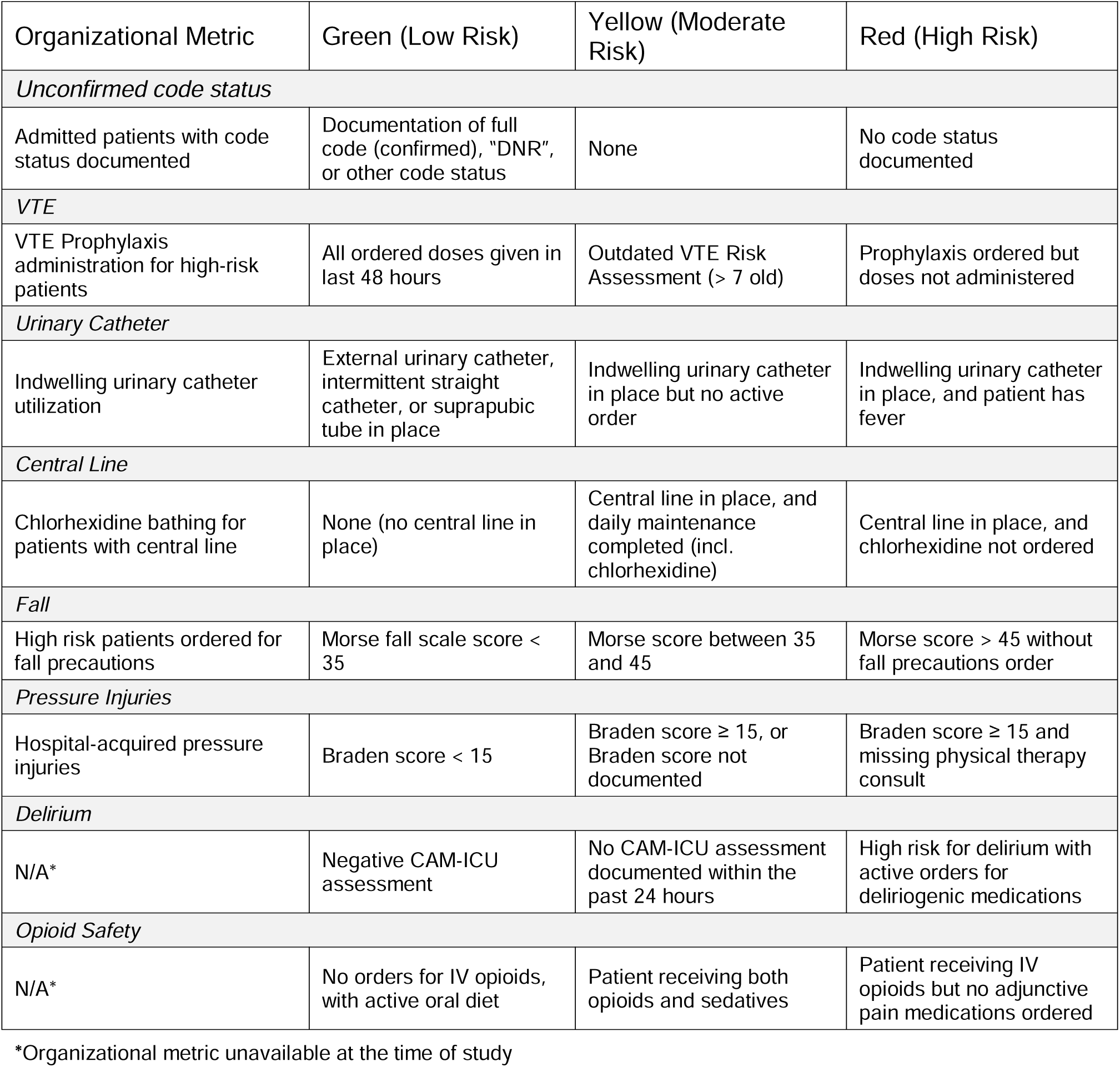
Organizational Metric and Examples of Dashboard Risk Categories, by Domain.

To support risk assessment and real-time action, the dashboard retrieved and consolidated structured EHR data corresponding to each safety category. These included orders, flowsheet items, encounter data, laboratory test and study results, and Epic-specific risk scores (e.g., readmission risk), accessed via an application programming interface (API). Using rule-based logic, the dashboard evaluated these data to flag patients as low risk (green), moderate risk (yellow), or high risk (red) for each harm domain. For each flagged risk state (**Table 2**), the dashboard provided real-time, patient-specific feedback on adherence to best practice guidelines and prompted clinicians to take targeted actions within the EHR (e.g., discontinue a device, complete an assessment, enter an order) to ensure compliance with key organizational metrics.

### Alignment with Local Organizational Initiatives and Leadership Engagement

We partnered with hospital quality leadership to ensure that the dashboard’s process metrics to align with existing organizational priorities. This alignment ensured that the dashboard would reinforce–rather than compete with–ongoing quality initiatives. We held stakeholder meetings and co-design sessions with clinical service chiefs, nurse managers, unit-based leadership, and frontline clinicians, focusing on refining the dashboard graphical user-interface (**Figure 2**) and identifying opportunities for workflow integration (such as during IDRs) to encourage adoption. We presented to the hospital leadership team, who endorsed the dashboard’s alignment with their local quality improvement initiatives.

### User Enhancements to Address Frontline Clinician Needs

To ensure alignment with the practical needs of frontline clinicians and institutionally defined metrics (**Table 2**), we configured the dashboard’s application logic and iteratively improved the graphical user interface (**Figure 2 and Supplemental Figure 1**). We conducted a series of design sessions with frontline clinician champions and stakeholders, focusing on usability by staff. All enhancements (**Supplemental Table 1**) were designed, tested, and deployed to encourage adoption and use by reducing cognitive burden, increasing relevance across roles, and promoting integration into daily workflows for frontline clinicians.

### Implementation and Integration into Clinical Workflows

The final version of the dashboard–featuring active domains related to code status, VTE, urinary catheters, central lines, falls, pressure injuries, delirium, and opioid safety–was launched in September 2019, the beginning of the post-implementation period. To support adoption, we implemented a multifaceted training and engagement strategy targeting all relevant clinical roles. Training sessions were delivered through dyad meetings, IDRs, teaching conferences, and Medicine Grand Rounds; and individual hands-on training for nursing staff, with research team members available to provide real-time support and gather feedback. Starting 11/11/19, we began producing weekly usage reports (**Supplemental Figure 2**), which included data on total logins, top users, and updates across units. These reports were shared with clinical leaders and unit staff to encourage ongoing engagement and recognize high-utilizing individuals and units.

### Training & Role-Specific Workflows

In collaboration with clinician champions and unit leadership, we implemented a workflow to support consistent dashboard use across interdisciplinary team members.^26^ Case managers and administrative staff led dashboard review during IDRs to discuss patients’ safety risks and progression of care.^26, 31^ Clinicians were prompted to address items such as unconfirmed code status and VTE prophylaxis, based on dashboard flags. Nurses were encouraged to review the dashboard for their assigned patients prior to IDRs. Physicians and APPs were advised to review the dashboard while admitting inpatients and writing notes. Role-specific focus areas also emerged: nocturnists focused on updating code status and confirming orders for fall precautions orders, while nurses concentrated on urinary catheter care, applying VTE prophylaxis, and delirium prevention efforts.

### Measures and Data Collection

Data for clinical outcomes, process measures, and implementation metrics were collected from the EHR via the Enterprise Data Warehouse (EDW); obtained from administrative reports and national quality measurement data and retrieved from the dashboard’s built-in performance auditing tool.

*Clinical outcomes* included 30-day post-discharge VTE; positive Confusion Assessment Measure (CAM) scores; hospital acquired pressure injuries, patient falls, CLABSIs, CAUTIs; and 30-day readmissions. *Process measures* corresponding to the dashboard’s safety domains included: code status documentation, use of mechanical and/or pharmacologic VTE prophylaxis, delirium risk assessments, ordering of chlorhexidine baths for patients with central lines, ordering urinalysis prior to urine cultures, documentation of Morse Fall Scale score, fall precaution orders for high risk patients, documentation of Braden Scale scores, and ordering physical therapy consults for patients at high risk of pressure injury. *Implementation metrics* included weekly counts of dashboard views and number of user clicks on dashboard items.

The primary outcome was the percentage of hospitalizations with at least one AE: post-discharge VTE, hospital acquired pressure injuries, falls with injury, CLABSIs, CAUTIs, opioid-related adverse drug events (naloxone administration, respiratory rate <10 breaths per minute, Richmond Agitation-Sedation Scale (RASS) score < -2 while receiving opioid therapy), use of mechanical restraints, positive CAM score. Severe hypoglycemia (glucose < 40) was tracked as a control measure.

Exploratory clinical outcomes included the rate of each individual HAC per 100 hospitalizations and adherence to associated institutionally-defined process measures. These process measures captured documentation of relevant orders or flowsheet entries in the EHR that aligned with hospital safety guidelines. Clinician engagement was assessed through the number of dashboard views per week during the study period.

### Statistical Analysis

All patients admitted to the study units during the pre-implementation and post-implementation periods were included in the analysis. The primary outcome–the rate of hospitalization with at least one AE–was compared between the post-vs. pre-implementation periods using Poisson regression to estimate incidence rate ratios (IRRs), with robust standard errors, adjusting for potential confounders including patient age, sex, race, median income by zip code, and Elixhauser comorbidity index

To evaluate temporal trends and the effect of the intervention over time, we conducted an interrupted time-series analysis using segmented regression by month, estimating baseline trend, change in level at implementation, and change in baseline trend between the pre- and post-implementation periods. Similar analyses were performed for each individual HAC and their corresponding process measures. Multivariable Poisson (for rates), logistic (for binary outcomes) and linear (for continuous process measures) regression models were similarly used to adjust for potential confounders including patient age, sex, race, median income by zip code, and Elixhauser comorbidity index.^32^ Run charts were created to illustrate exploratory measures with significant changes in the post-implementation period.

All models, both unadjusted and adjusted, incorporated general estimating equations (GEE) to account for clustering by hospital unit. As is standard for interrupted time series, we considered significant changes in slope or y-intercept to be statistically significant. Given the exploratory nature of the secondary outcomes, we did not adjust for multiple testing. All analyses were conducted using SAS 9.1 (SAS Institute, Cary, NC).

## RESULTS

A total of 9,023 hospitalizations occurred during the pre-implementation (n=4,640) and post-implementation (n=4383) periods. Demographic characteristics (**Table 3**) were largely comparable between the two periods, with most variables differing by <5% in absolute terms. Mean LOS was slightly longer during the post-implementation (5.07 days vs 5.44 days) , while median LOS remained unchanged (4.00 days). Run charts for exploratory measures with significant changes are illustrated in **Figure 3**.

**Figure 3.**
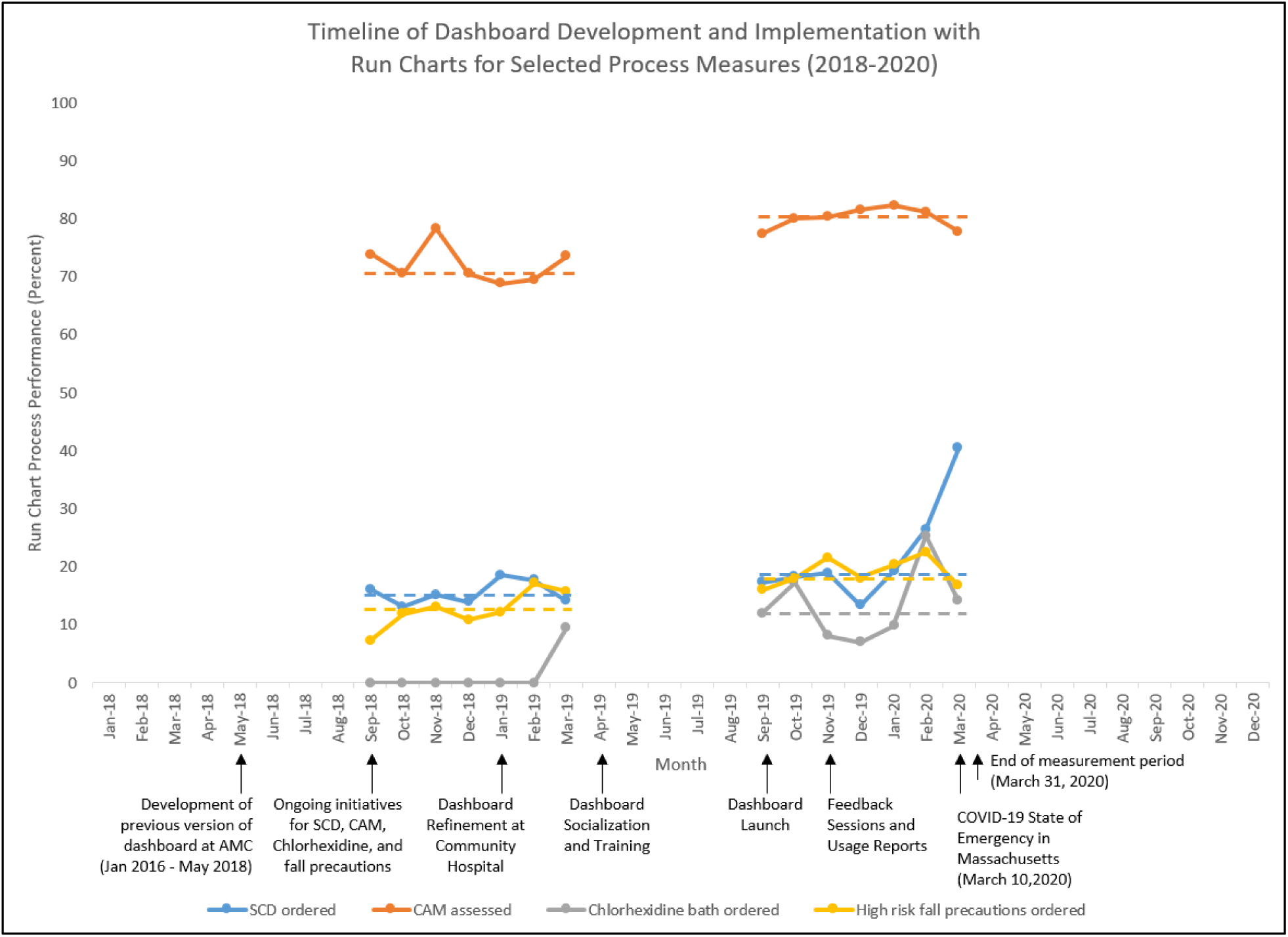
Timeline of Dashboard Development and Implementation with Run Charts for Selected Process Measures (2018-2020). Exploratory outcomes with significant post-implementation changes from **Table 4** are displayed as run charts, with median baseline performance as dotted lines.

**Table 3.** Baseline characteristics of hospitalizations.

| Characteristic | Pre, n=4640 | Post, n=4383 |
| --- | --- | --- |
| **No. unique patients** | 3678 | 3528 |
| **Age in years–mean (SD)** | 65.5 (18.8) | 65.5 (18.2) |
| **Female gender–no. (%)** | 2649 (57.1) | 2414 (55.1) |
| **Length of Stay - days** |  |  |
| Mean (median) | 5.07 (4.00) | 5.44 (4.00) |
| **Race–no. (%)** |  |  |
| White | 2909 (62.7) | 2672 (61.0) |
| Black | 711 (15.3) | 793 (18.1) |
| Hispanic | 715 (15.4) | 601 (13.7) |
| Other/Missing | 305 (6.6) | 317 (7.2) |
| **Ethnicity – no. (%)** |  |  |
| Non-Hispanic | 3389 (73.1) | 3408 (77.8) |
| Hispanic | 681 (14.7) | 585 (13.4) |
| Unavailable | 561 (12.1) | 382 (8.7) |
| Declined | 8 (0.2) | 6 (0.1) |
| **Marital Status – no. (%)** |  |  |
| Partnered | 1660 (35.8) | 1516 (34.6) |
| Single | 2843 (61.3) | 2736 (62.4) |
| Unknown | 137 (3.0) | 131 (3.0) |
| **Primary language English – no. (%)** | 3932 (84.7) | 3757 (85.7) |
| **Education – no. (%)** |  |  |
| Less than high school | 664 (14.3) | 603 (13.8) |
| Graduated high school | 1944 (41.9) | 1924 (43.9) |
| Graduated college/Higher Education | 1312 (28.3) | 1242 (28.3) |
| Unknown | 720 (15.5) | 614 (14.0) |
| **Socioeconomic status (median income by zip code) – no. (%)** |  |  |
| Less than or equal to \$47,000 | 550 (11.9) | 618 (14.1) |
| \$47,001 to \$63,000 | 1503 (32.4) | 1421 (32.4) |
| Greater than \$63,000 | 2564 (55.3) | 2318 (52.9) |
| Missing | 23 (0.5) | 26 (0.6) |
| **Insurance status – no. (%)** |  |  |
| Private | 1727 (37.2) | 1638 (37.4) |
| Medicaid | 522 (11.3) | 522 (11.9) |
| Medicare | 2254 (48.6) | 2130 (48.6) |
| Other | 137 (3.0) | 93 (2.1) |
| **Primary care physician – no. (%)** |  |  |
| Network | 2359 (50.8) | 2273 (51.9) |
| Non-network | 2281 (49.2) | 2110 (48.1) |
| **Elixhauser comorbidity index – no. (%)** |  |  |
| Less than or equal to 0 | 1205 (26.0) | 942 (21.5) |
| 1 to 5 | 1054 (22.7) | 943 (21.5) |
| 6 to 10 | 857 (18.5) | 838 (19.1) |
| 11 or more | 1524 (32.8) | 1660 (37.9) |
| **Study Unit – no. (%)** |  |  |
| Unit 1 (6N) | 1398 (30.1) | 1277 (29.1) |
| Unit 2 (6S) | 1736 (37.4) | 1506 (34.4) |
| Unit 3 (7N) | 1506 (32.5) | 1600 (36.5) |

**Table 4.**
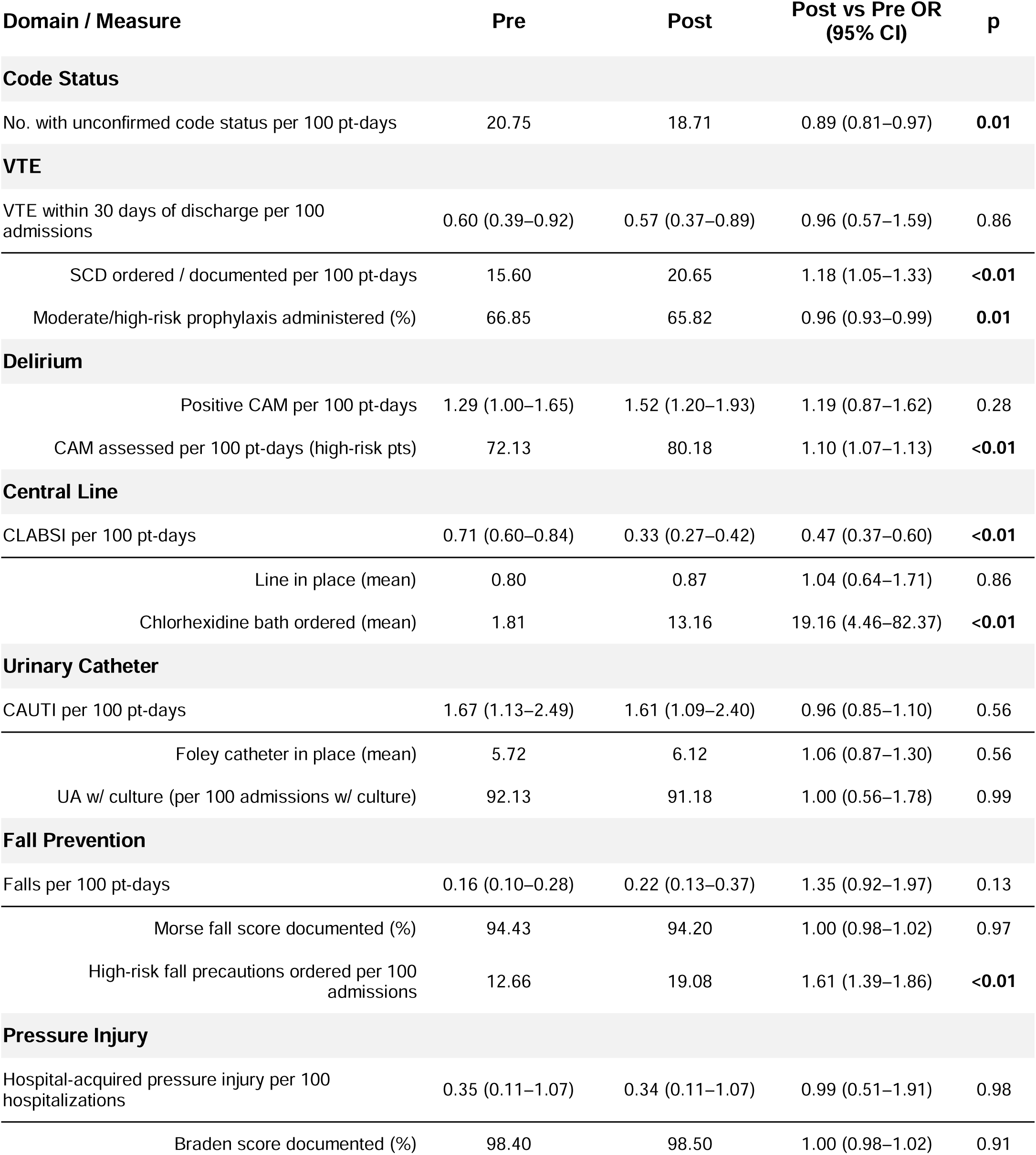
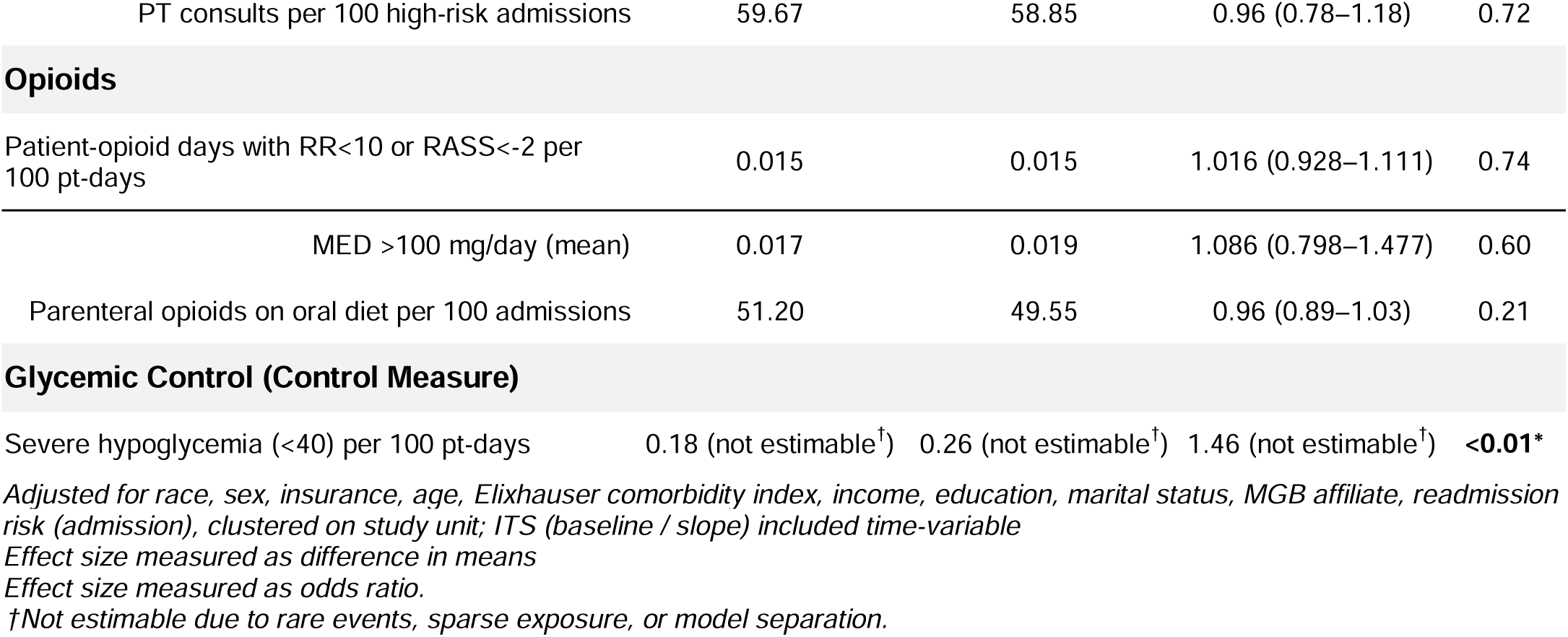
Exploratory Outcomes: Domain Specific In-Hospital Outcome & Process Measures.

The percentage of hospitalizations with an AE was similar in the pre-implementation period (23.15%; 95% CI, 20.96–25.29) and post-implementation period (22.97%; 95% CI, 20.86–25.23), with no significant change in the adjusted incidence rate ratio (IRR, 0.99; 95% CI, 0.90–1.10; p=0.93), nor evidence of a significant immediate level change (intercept change IRR, 0.94; 95% CI, 0.68–1.31; p=0.72) or change in slope over time (slope change IRR, 0.99; 95% CI, 0.94–1.04; p=0.72).

Analysis of individual HAC outcomes showed a significant pre-post reduction in CLABSI per 100 patient-days (aOR 0.47, 95% CI [0.37-0.60]) (**Table 4**). Interrupted time series analysis showed a significant decrease in slope for post-discharge VTE (aOR 0.66, 95% CI [0.48,0.91], p=0.01) with a non-significant change in y-intercept; and a decrease in slope with a non-significant change in y-intercept for hospital-acquired pressure injury (aOR 0.69, 95% CI [0.48, 1.00], p=0.05) (**Supplemental Table 2**). Several process measures demonstrated significant improvement from the pre-to post-implementation periods (**Table 4**, **Figure 3** and **Supplemental Table 2)**. Hospitalizations with unconfirmed code status decreased significantly (OR 0.89, 95% CI [0.81, 0.97], p=0.01), along with significant decreases in both slope and y-intercept. Documentation of sequential compression device (SCD) use for DVT prophylaxis increased (OR 1.18, 95% CI [1.05, 1.33], p<0.01) and was associated with a significant positive slope change. Compliance with CAM assessments among high-risk patients improved significantly (OR 1.10, 95% CI [1.07, 1.13], p<0.01). Orders for chlorhexidine bathing increased (OR 19.16, 95% CI [4.46, 82.37], p<0.01), and orders for fall precaution in high-risk patients increased (OR 1.61, 95% CI [1.39, 1.86], p<0.01). One measure worsened: the administration of medical DVT prophylaxis in moderate or high-risk patients (OR 0.96 [0.93, 0.99], p=0.01).

Severe hypoglycemia events per 100 patient-days—an outcome not targeted by the intervention—were rare but increased in the postimplementation period (OR 1.46). Due to sparse events, confidence intervals could not be reliably estimated; however, a statistically significant change in slope was observed.

Dashboard utilization increased steadily over the post-implementation period (**Supplemental Figure 3),** rising from 10 views per week in the first week to 80 views per week by week 16. Weekly usage reports indicated consistent engagement across clinical roles, with variation by unit and provider type. Dashboard use dropped sharply to 20 views per week by week 17, coinciding with the onset of the COVID-19 pandemic in March 2020 and the suspension of in-person IDRs due to mandatary physical distancing measures due to the state of emergency declared on March 10, 2020. During the study period, front-line clinicians using the dashboard commented on their unit being Foley-free for four days (hospitalist), using the dashboard to facilitate engagement (nurse director), and competing to become the top dashboard user (nurses).

## DISCUSSION

In this quality improvement study, we evaluated a customized, EHR-integrated dashboard designed to support real-time identification of risks for hospital-acquired conditions among general medicine inpatients in an academic community hospital. The dashboard interface was iteratively refined in collaboration with hospital leadership, unit-based clinicians, and quality and safety teams to align with institutional priorities and frontline workflows. Although the primary outcome—the rate of hospitalizations with at least one adverse event—did not significantly improve following implementation, several related outcome and process measures demonstrated meaningful improvements in exploratory analyses. Dashboard utilization also increased and was sustained throughout the implementation period until the onset of the COVID-19 pandemic.

Significant process improvements were observed in documentation and ordering practices across key safety domains, including code status confirmation, mechanical venous thromboembolism prophylaxis, delirium screening, fall precautions, and chlorhexidine bathing. Hospitalizations with unconfirmed code status—a measure aimed at preventing harm from unnecessary aggressive resuscitation efforts (e.g., intubation, chest compressions)—decreased significantly, likely reflecting consistent dashboard use during interdisciplinary rounds (IDRs), where case managers prompted teams to address outstanding documentation gaps. Most other improved process measures were largely nurse-driven, reflecting nurses’ consistent presence and engagement with the initiative (as opposed to residents and attendings, who move between different teams and also work at BWH, the main teaching hospital). Despite improvements in process measures, the rate of CLABSI was the only outcome measure with a notable improvement. These findings are consistent with the Donabedian model, which emphasizes that improvements in care processes—particularly those closely linked to desired outcomes—may serve as early indicators of quality improvement success, even when downstream clinical outcomes do not change in the short term.^33^

The dashboard’s success in improving process adherence was likely driven by its user-centered design within our patient safety research program.^21–28, 34^ By engaging frontline clinicians early in the design process, tailoring content to clinical roles, and linking alerts directly to actionable EHR functions, the dashboard was better aligned with clinicians’ workflows, which may explain its acceptability despite concerns about cognitive burden. The rise in dashboard use over time, followed by a sharp decline coinciding with the onset of the COVID-19 pandemic and the suspension of in-person IDRs, underscores the central role of contextual factors in sustaining engagement. Dashboard use appeared most effective when embedded within established daily workflows rather than functioning as a stand-alone tool. Notably, many users perceived the dashboard positively and continued to use it after the conclusion of the study, suggesting durable integration into routine clinical practice. Nevertheless, several measures did not improve, particularly those involving more complex processes such as opioid management and early discontinuation of urinary catheter use. Improvement was greatest in the domains requiring one simple action to be in compliance, e.g., documenting an action such as code status, CAM score, SCD use, or ordering fall precautions.

While few studies have evaluated real-time, EHR-integrated dashboards focused on inpatient quality and safety, our work offers a pragmatic model for measure selection, dashboard design, and implementation. This study is among the first to demonstrate the potential for improving individual processes by aligning institutional priorities with clinician end-user needs within routine clinical workflows. Our approach—codesigned with end users, responsive to clinical context, and integrated into existing EHR infrastructure—may be generalizable to other settings seeking to enhance quality and safety performance using real-time data. Future iterations of the dashboard could be further refined to strengthen its impact on preventing HACs and improving other quality outcomes, such as simplifying its messages and actions to be taken. Further actions may be needed to fully integrate dashboard use into routine care on a busy inpatient service.

While our study has notable strengths, including a collaborative design process and the integration of real-time EHR data to provide actionable insights about HAC risk, several limitations should be considered. First, as a single-site pre-post study, our findings have limited generalizability and may be subject to confounding from concurrent initiatives and external factors, including disruptions related to the COVID-19 pandemic (March 10, 2020), which overlapped the measurement period during the final three weeks (March 31, 2020). Notably, chlorhexidine bathing was initiated at the end of the baseline period, which confounds the effect of the dashboard on this process (**Figure 3**). While it is difficult to separate the effects of the dashboard from other co-interventions, staff reported positive experiences with using the dashboard to improve their performance on care processes, such as removing foley catheters. Second, most observed improvements involved documentation-dependent processes, and the dashboard’s impact on lower-incidence clinical outcomes (e.g., falls with injury or infection rates) was less clear. Third, usage metrics were measured at the unit level, limiting more granular analysis by clinician role or specialty. Fourth, while we did not analyze end-clinician user experience formally, the previous version of the dashboard underwent rigorous human-centered design methods – the current version relied on rapid iterative enhancements aligned with workflows and were informally positively received.^22–24^ Finally, the dashboard was custom developed within our patient safety research program, and substantial effort would be required to replicate and scale this functionality within commercial EHR platforms at other sites.

In summary, this study demonstrates that an EHR-integrated dashboard can support quality improvement efforts across multiple HAC-related domains. A user-centered design process and alignment with institutional priorities facilitated-adoption, improved process adherence, and supported sustained engagement by frontline clinicians. Future work should explore further attempts to integrate dashboard use within clinical care, broader implementation across services, refinement of outcome measures, and longer-term effects on patient safety outcomes.

## Data Availability

All data produced in the present study are available upon reasonable request to the authors

## Contributorship

All authors have contributed sufficiently and meaningfully to the conception, design, and conduct of the study; data acquisition, analysis, and interpretation; and/or drafting, editing, and revising the manuscript.

## SUPPLEMENTAL MATERIALS

**Supplemental Figure 1:**
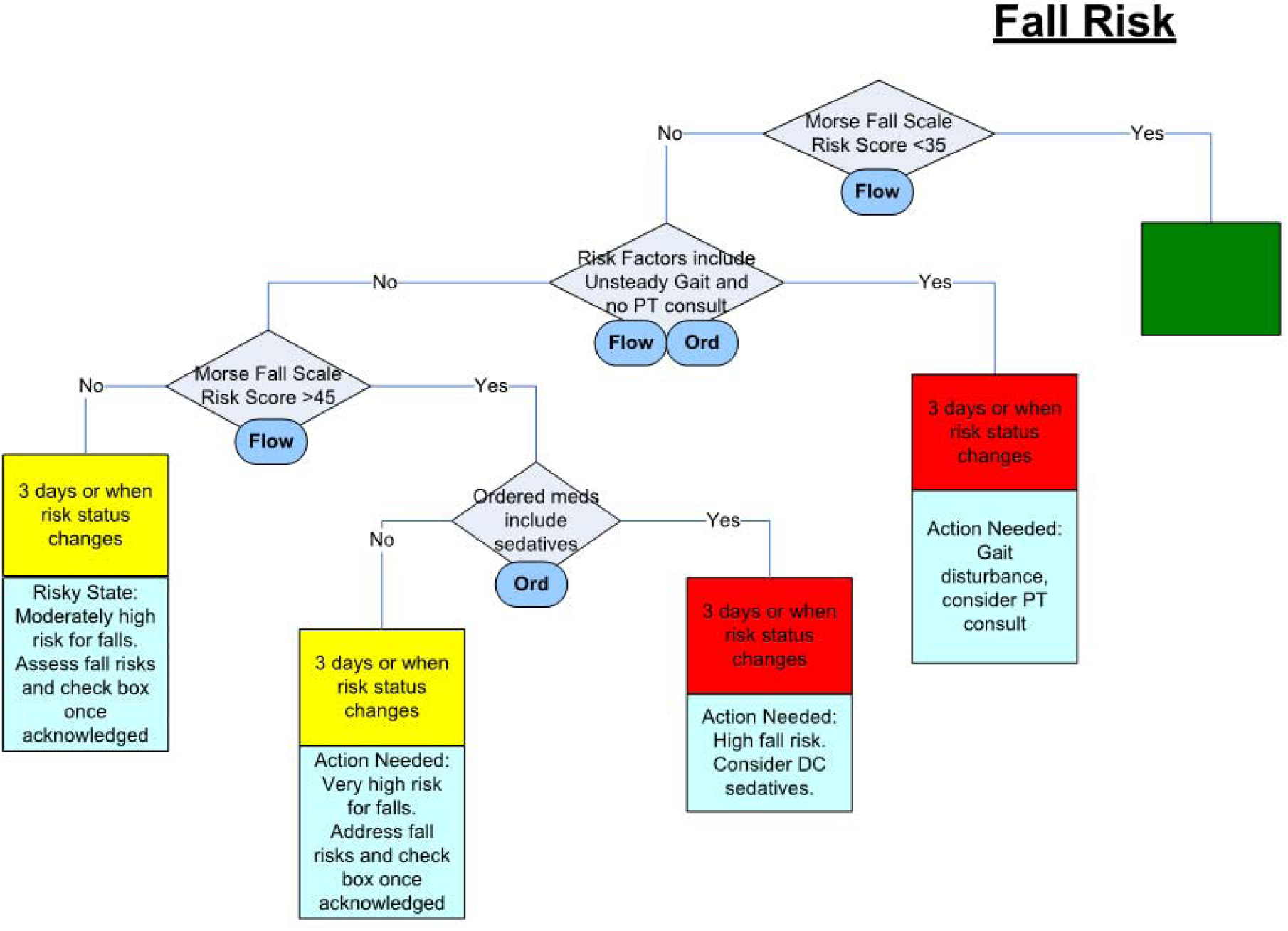
Example of Dashboard Logic for Falls Prevention

**Supplemental Figure 2:**
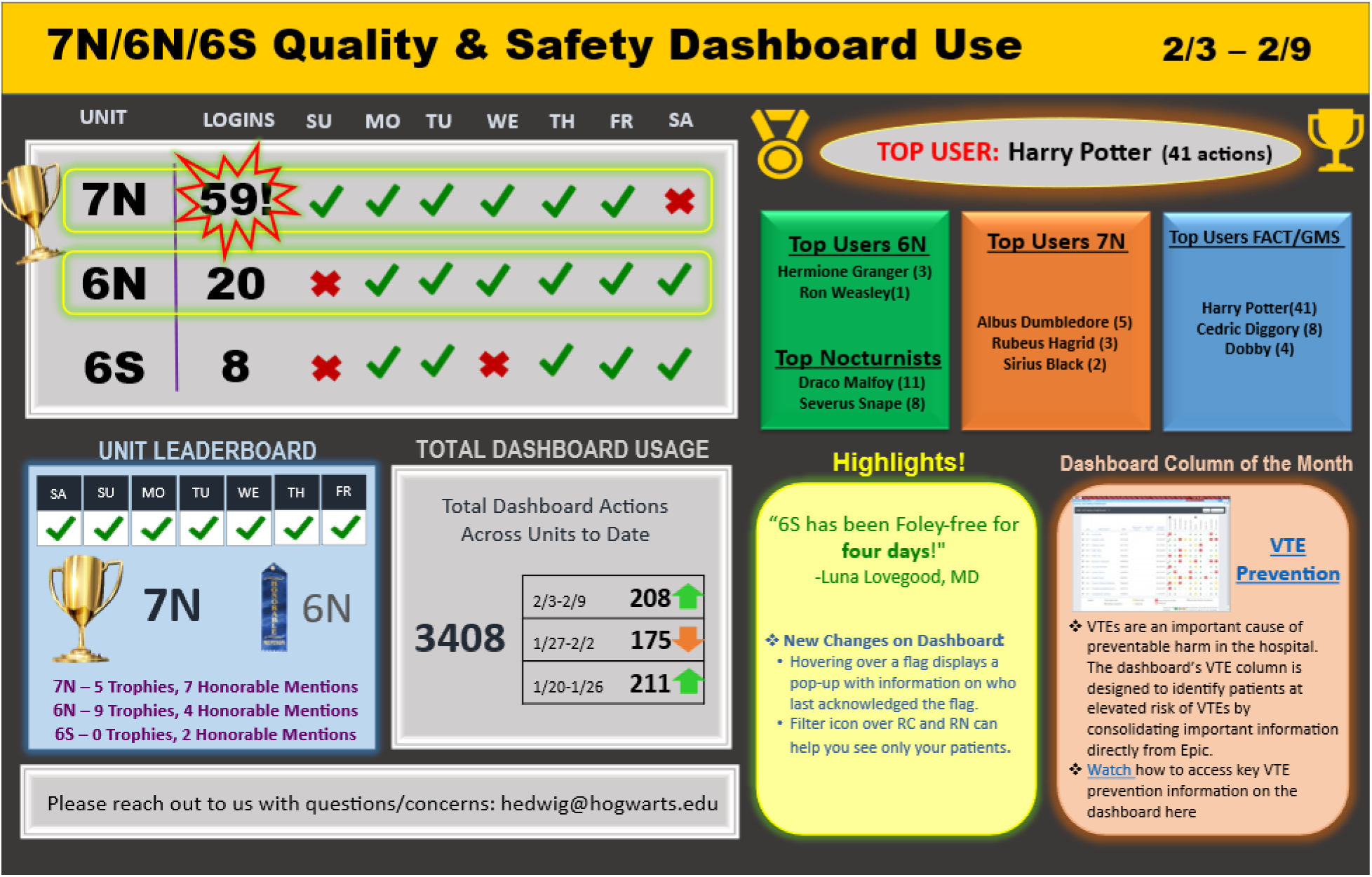
Weekly usage reports

**Supplemental Figure 3.**
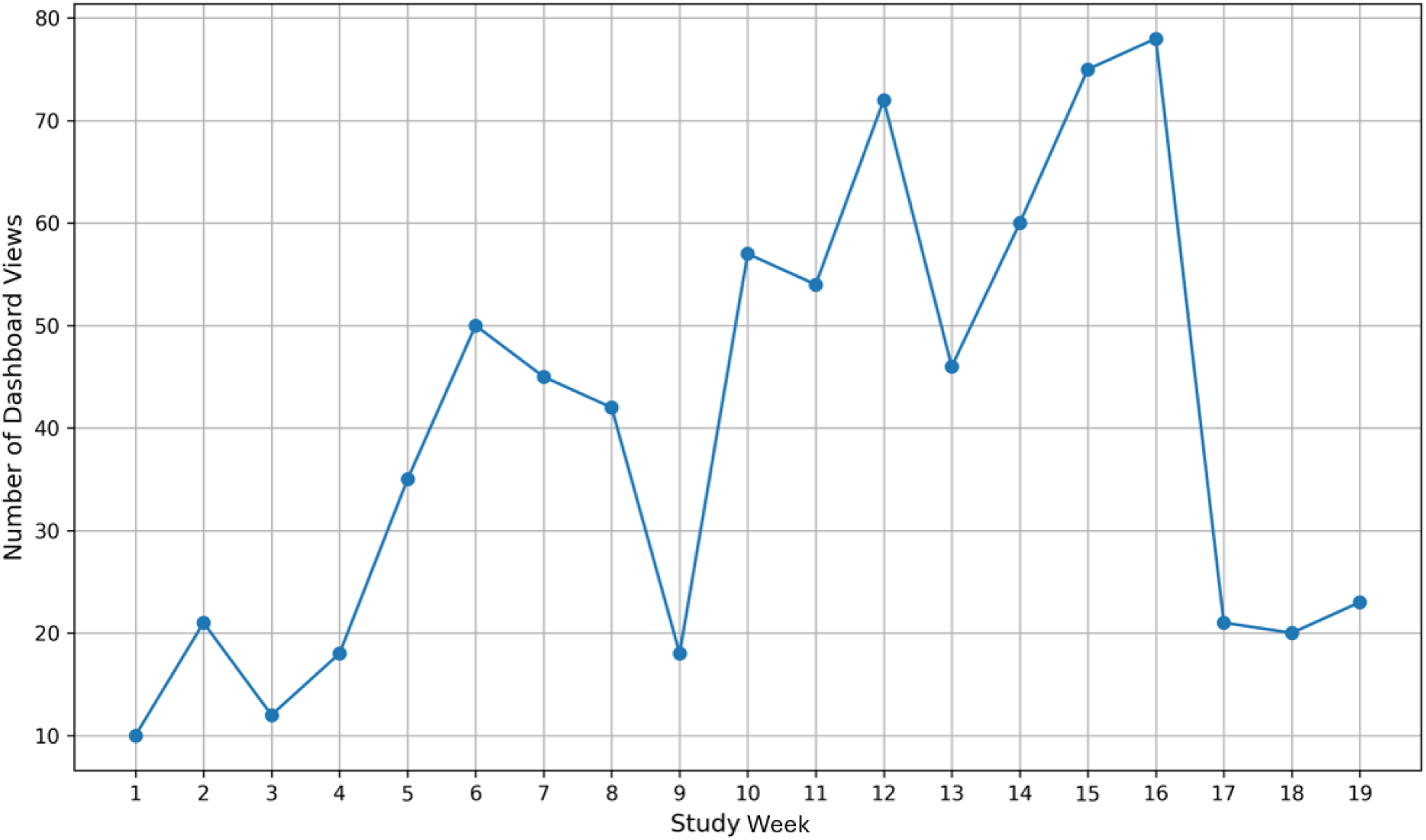
Trends in Dashboard Utilization by Clinicians Post-Implementation

**Supplemental Table 1.** Enhancements required to encourage adoption and use by front line clinicians.

| Enhancement | Description | Perceived benefits for clinicians |
| --- | --- | --- |
| Unit and team lists | View patient list for specific user roles. | Service-based staff (physicians, APPs) prefer patient lists arranged by care team, and unit-based staff (RNs) prefer lists arranged by patient location |
| Patient chart view | Access all risk flags from within the patient's chart | Offers ability to view the dashboard's decision support directly within the patient chart in the EHR |
| Actionable hyperlinks | Users can take immediate actions from dashboard flags (e.g., place orders, update flowsheets) | Offers ability to act upon decision support from the patient's chart in the EHR to document assessments or place orders (e.g., order urinary catheter removal) |
| Sort | Sort by bed number, patient name, MRN, responding clinician, or registered nurse. | Eased the burden of locating patients in a unit or team view |
| Notifications | Configurable notification badge<br>Displays new flag updates in last 8 hours | Provided quick way of identifying new flags over certain time interval |
| Logic definitions | Clickable information buttons<br>Define risks and recommended actions for each domain | Clearly explain for users why certain colored flags would appear |
| Defer button | "Defer" button to delay or silence risk flags for predefined time intervals | Reduce alarm fatigue by deferring review of high-risk flags for certain exclusions, e.g., use of central line in patient with poor access, holding VTE prophylaxis given active bleeding, etc) |

**Supplemental Table 2.** Interrupted Time Series Effects: Baseline Trend, Intercept Change, and Slope Change (Exploratory Outcomes).

| Domain / Measure | Intercept Change aOR<br>(95% CI) | p | Slope Change aOR<br>(95% CI) | p |
| --- | --- | --- | --- | --- |
| **Code Status** |  |  |  |  |
| No. with unconfirmed code status per 100 pt-days | 0.39 (0.28, 0.53) | <0.01 | 0.83 (0.79, 0.87) | <0.01 |
| **VTE** |  |  |  |  |
| VTE within 30 days of discharge per 100 admissions | 0.71 (0.16, 3.22) | 0.66 | 0.66 (0.48, 0.91) | 0.01 |
| SCD ordered, documented use per 100 pt-days | 0.91 (0.61, 1.35) | 0.64 | 1.12 (1.06, 1.19) | <0.01 |
| Moderate or high risk, medical prophylaxis administered | 0.96 (0.86, 1.07) | 0.44 | 0.99 (0.97, 1.00) | 0.18 |
| **Delirium** |  |  |  |  |
| Positive CAM per 100 pt-days | 0.67 (0.23, 1.92) | 0.45 | 0.91 (0.78, 1.06) | 0.24 |
| CAM assessed in highrisk patients per 100 pt-days | 1.06 (0.96, 1.18) | 0.25 | 1.01 (0.99, 1.02) | 0.29 |
| **Central Line** |  |  |  |  |
| CLABSI per 100 pt-days | 0.66 (0.30, 1.47) | 0.31 | 1.00 (0.88, 1.14) | 0.99 |
| Line in place (mean) | 0.92 (0.17, 4.97) | 0.93 | 1.10 (0.85, 1.44) | 0.46 |
| Chlorhexidine bath ordered (mean) | Not estimable<sup>†</sup> | - | 0.05 (0.0005, 5.10) | 0.20 |
| **Urinary Catheter** |  |  |  |  |
| CAUTI per 100 pt-days | 0.72 (0.42, 1.24) | 0.22 | 1.00 (0.92, 1.09) | 0.99 |
| Foley catheter in place (mean) | 0.92 (0.47, 1.80) | 0.82 | 1.01 (0.91, 1.11) | 0.83 |
| Urine culture with UA ≤72h per 100 admissions with urine culture | 0.55 (0.04, 7.88) | 0.65 | 1.08 (0.65, 1.80) | 0.77 |
| **Fall Prevention** |  |  |  |  |
| Falls per 100 pt-days | 0.54 (0.12, 2.42) | 0.42 | 1.11 (0.89, 1.37) | 0.36 |
| Morse fall score documented (mean) | 1.00 (0.94, 1.07) | 0.897 | 0.998 (0.99, 1.01) | 0.74 |
| High-risk (Morse >45), fall precautions ordered per 100 admissions | 0.76 (0.46, 1.25) | 0.28 | 0.98 (0.91, 1.05) | 0.52 |
| **Pressure Injury** |  |  |  |  |
| Hospital-acquired pressure injury per 100 hospitalizations | 0.36 (0.04, 2.91) | 0.34 | 0.69 (0.48, 1.00) | 0.05 |
| Braden score documented (mean) per 100 admissions | 1.00 (0.94, 1.07) | 0.97 | 1.00 (0.99, 1.01) | 0.99 |

| Domain / Measure | Intercept Change aOR<br>(95% CI) | p | Slope Change aOR<br>(95% CI) | p |
| --- | --- | --- | --- | --- |
| PT consults per 100 high-risk admissions | 0.73 (0.34, 1.57) | 0.42 | 0.96 (0.86, 1.07) | 0.45 |
| **Opioids** |  |  |  |  |
| Patient-opioid days with RR<10 or RASS<-2 per 100 pt-days | 0.77 (0.57, 1.03) | 0.08 | 1.00 (0.96, 1.05) | 0.99 |
| MED >100 mg/day (mean) | 0.86 (0.31, 2.36) | 0.77 | 1.04 (0.89, 1.21) | 0.61 |
| Parenteral opioids on oral diet per 100 admissions | 1.12 (0.90, 1.42) | 0.34 | 0.99 (0.96, 1.03) | 0.64 |
| Severe hypoglycemia (<40) per 100 pt-days | 4.14 (0.22, 79.00) | 0.34 | 1.19 (not estimable<sup>†</sup>) | **<0.01** |

*Adjusted for race, sex, insurance, age. Elixhauser comorbidity index, income, education, marital status, MGB affiliate, readmission risk (admission), clustered on study unit; ITS (baseline / slope) included time-variable*
*Effect size measured as difference in means*
*Effect size measured as odds ratio*
*†Not estimable due to rare events, sparse exposure, or model separation.*

